# The central nervous system’s proteogenomic and spatial imprint upon systemic viral infections with SARS-CoV-2

**DOI:** 10.1101/2023.01.16.22283804

**Authors:** Josefine Radke, Jenny Meinhardt, Tom Aschman, Robert Lorenz Chua, Vadim Farztdinov, Sören Lukkassen, Foo Wei Ten, Ekaterina Friebel, Naveed Ishaque, Jonas Franz, Valerie Helena Huhle, Ronja Mothes, Kristin Peters, Carolina Thomas, Simon Streit, Regina von Manitius, Péter Körtvélyessy, Stefan Vielhaber, Dirk Reinhold, Anja Hauser, Anja Osterloh, Philipp Enghard, Jana Ihlow, Sefer Elezkurtaj, David Horst, Florian Kurth, Marcel A. Müller, Nils C. Gassen, Julia Schneider, Katharina Jechow, Bernd Timmermann, Camila Fernandez-Zapata, Chotima Böttcher, Werner Stenzel, Emanuel Wyler, Victor Corman, Christine Stadelmann-Nessler, Markus Ralser, Roland Eils, Frank L. Heppner, Michael Mülleder, Christian Conrad, Helena Radbruch

## Abstract

In COVID-19 neurological alterations are noticed during the systemic viral infection. Various pathophysiological mechanisms on the central nervous system (CNS) have been suggested in the past two years, including the viral neurotropism hypothesis. Nevertheless, neurological complications can also occur independent of neurotropism and at different stages of the disease and may be persistent.

Previous autopsy studies of the CNS from patients with severe COVID-19 show infiltration of macrophages and T lymphocytes, especially in the perivascular regions as well as pronounced microglial activation, but without signs of viral encephalitis.

However, there is an ongoing debate about long-term changes and cytotoxic effects in the CNS due to the systemic inflammation.

Here, we show the brain-specific host response during and after COVID-19. We profile single-nucleus transcriptomes and proteomes of brainstem tissue from deceased COVID-19 patients who underwent rapid autopsy. We detect a disease phase-dependent inflammatory type-I interferon response in acute COVID-19 cases. Integrating single-nucleus RNA sequencing and spatial transcriptomics, we could localize two patterns of reaction to severe systemic inflammation. One neuronal with direct focus on cranial nerve nuclei and one diffusely affecting the whole brainstem, the latter reflecting a bystander effect that spreads throughout the vascular unit and alters the transcriptional state of oligodendrocytes, microglia and astrocytes.

Our results indicate that even without persistence of SARS-CoV-2 in the CNS, the tissue activates highly protective mechanisms, which also cause functional disturbances that may explain the neurological symptoms of COVID-19, triggered by strong systemic type-I IFN signatures in the periphery.

## Introduction

COVID-19 can be used as an example to study cerebral alterations during a systemic viral infection as many of the inflammatory changes noticed during COVID-19 are not SARS-CoV-2-specific. On the one side, neurological complications^1–6^ can occur independent of viral neurotropism and at different stages of the disease and may be persistent^7^. On the other hand, various pathophysiological mechanisms of COVID-19-associated effects on the CNS have been suggested in the past two years, including the neurotropism hypothesis. This hypothesis is based on the detection of viral RNA, while active replication of SARS-CoV-2 or intact viral particles have not yet been shown in the CNS^8,9^. Nevertheless, existing histological and molecular data implicate immune activation and inflammation within the CNS as well as dysregulation of CNS-specific cells primarily caused by indirect systemic effects^8,10^. Previous autopsy studies of the CNS from patients with severe COVID-19 show infiltration of macrophages, CD8+ T lymphocytes especially in the perivascular regions, and pronounced microglial activation^11–14^, however, without signs of viral encephalitis with lytic, infected cells. Single-cell RNA sequencing as well as flow cytometric studies of tissue and blood cells have revealed dysregulated myeloid cell responses in severe COVID-19^15^ and increased numbers of dedifferentiated monocytes and unconventionally activated T cells in the blood^16^. Additionally, the antiviral IFN type I signature is highly enriched in the CSF^17^ and blood^16,18^ of these individuals.

To elucidate the cellular immune-mediated mechanisms, cell composition, and host responses within the brain at molecular and topographic level, we analyzed the proteo-transcriptomic landscape and spatial architecture at different disease stages during and after acute COVID-19 in the brainstem. So far, there is no robust proof of SARS-CoV-2 entering the brain along the cranial nerves, reaching cardio-respiratory control centers in the brainstem^7^. However, there is neurophysiological evidence of SARS-CoV-2-related brainstem involvement in severe COVID-19 patients, especially at the medullary level^19^. In addition, 18F-FDG PET showed hypometabolism in the brainstem and cerebellum, which is associated with the occurrence of certain symptoms (hyposmia/anosmia, memory/cognitive impairment, pain and insomnia). We included controls with similar frequencies of multi-organ failure and bacterial superinfections as these are common comorbidities in hospitalized severe COVID-19 patients. Together, our findings show an engagement of the brainstem due to the systemic infection in COVID-19 beyond the acute phase, which could explain (prolonged) neurological symptoms such as impaired awakeness, pain or cardio-respiratory control, and provide insights into the neuroimmunology of other viral infections as well as systemic “interferonopathies” affecting the CNS.

## Results

### Sample characterization for proteogenomic postmortem analysis

In order to determine proteo-transcriptomic perturbations in the central nervous system (CNS) during systemic infection we studied patients with COVID-19 in different disease states. We analyzed fresh-frozen preserved postmortem tissue samples of cerebellum, olfactory bulb, and brainstem; the latter two regions represent potential viral entry routes to the CNS. The olfactory bulb is in close anatomical proximity to the olfactory mucosa (the primary site of infection) and the brainstem exerts specific regulatory functions of the cardio-respiratory system and contains multiple cranial nerve nuclei (e.g. vagal and hypoglossal nerve nuclei). The cerebellum served as internal CNS control with no direct synaptic connections to the olfactory system or cranial nerve nuclei. Lung and olfactory mucosa were taken as controls for primary infection sites.

First, tissue samples of the different anatomical sites were screened for viral RNA load by RT-qPCR. Lung and olfactory mucosa demonstrated the highest levels of SARS-CoV-2 RNA, while only low levels of viral RNA were detected in samples obtained from the olfactory bulb, brainstem and cerebellum. Viral load in lung and mucosa inversely correlated with disease duration, i.e. lower levels were found in late cases (**Supplementary Table 1**). Using immunohistochemistry, we detected no SARS-CoV-2 nucleocapsid protein in PCR-positive CNS samples (see **Supplementary Material** at Zenodo (doi: 10.5281/zenodo.7524299)) As these samples all showed very low RNA levels (below the number of copies expected for a single infected cell^9^), we assumed contamination or RNA debris in these samples rather than infection of CNS cells.

### Whole-tissue proteomes distinguish acute from late COVID-19 in the CNS

Next, we performed proteomic profiling of cryopreserved autopsy tissue samples from the olfactory bulb (n = 18), cerebellum (n = 21), brainstem (n = 22), and olfactory mucosa (n = 21). In total, around 4500 different proteins were quantified in all tissue types. The numbers of quantified proteins in each brain region were different, ranging from approximately 3000 proteins (olfactory mucosa) to 4000 proteins (cerebellum).

We divided the COVID-19 cohort into acute and late cases according to disease duration (14 days post infection as cut-off; **Supplementary Table 1**) due to marked differences in lung pathology seen in these patients^20^. Therefore, we first performed post hoc principal component analysis (PCA) of the differentially expressed proteins in the different comparisons (acute vs. control and late vs. control) of the brainstem. Principal components (PC) represent directions of the data that explain the maximum amount of variance. The first PC describes the largest contribution. The score plot shows a clustering of the brainstem samples in acute, late and control groups in both comparisons. We further analyzed the features contributing most to the first two principal components (PC) and clinical factors such as age, highest procalcitonin (PCT) levels before death, viral load, disease duration, and sex with the loading plot. We only detected a small contribution of the sample distribution in this analysis by age into the direction of “acute”. The second PC was influenced by clinical/demographic factors: highest PCT, viral load, disease duration, and male sex. Of note, PCT is a surrogate marker for bacterial superinfection (**Supplementary Figure 1**). Because of the results of the PCA in the brainstem, we continued our further analysis by separating acute and late COVID-19 donors into two distinct groups.

To identify significantly enriched pathways, we applied gene set enrichment analysis (GSEA) of the proteome dataset, in which the brainstem samples demonstrated a strong upregulation of pathways connected to COVID-19 such as: ‘Coronavirus disease - COVID-19*’* (ID: hsa05171), *‘*Interferon signaling’ (ID: R-HSA-913531), ‘Interferon gamma signaling’ (ID: R-HSA-877300), or ‘Antiviral mechanism by IFN− stimulated genes’ (R− HSA− 1169410) among others in the acute disease phase (**Figure 1 a**, highlighted by black boxes). Comparing olfactory bulb, cerebellum, and brainstem, the cerebellum showed the strongest upregulation of complement and coagulation cascades (ID: hsa04610), but less upregulation of interferon-related and virus associated pathways. In ‘late*’* COVID-19, Interferon-associated pathways were clearly less upregulated and pathways directly connected to COVID-19 (ID: hsa05171) or viral translation (e.g. *‘*Viral mRNA translation’ ID: R-HAS-192823)) were downregulated which again underlined our rationale to separate the COVID-19 cohort into ‘acute*’* and ‘late*’* (**Figure 1 a**).

**Figure 1:**
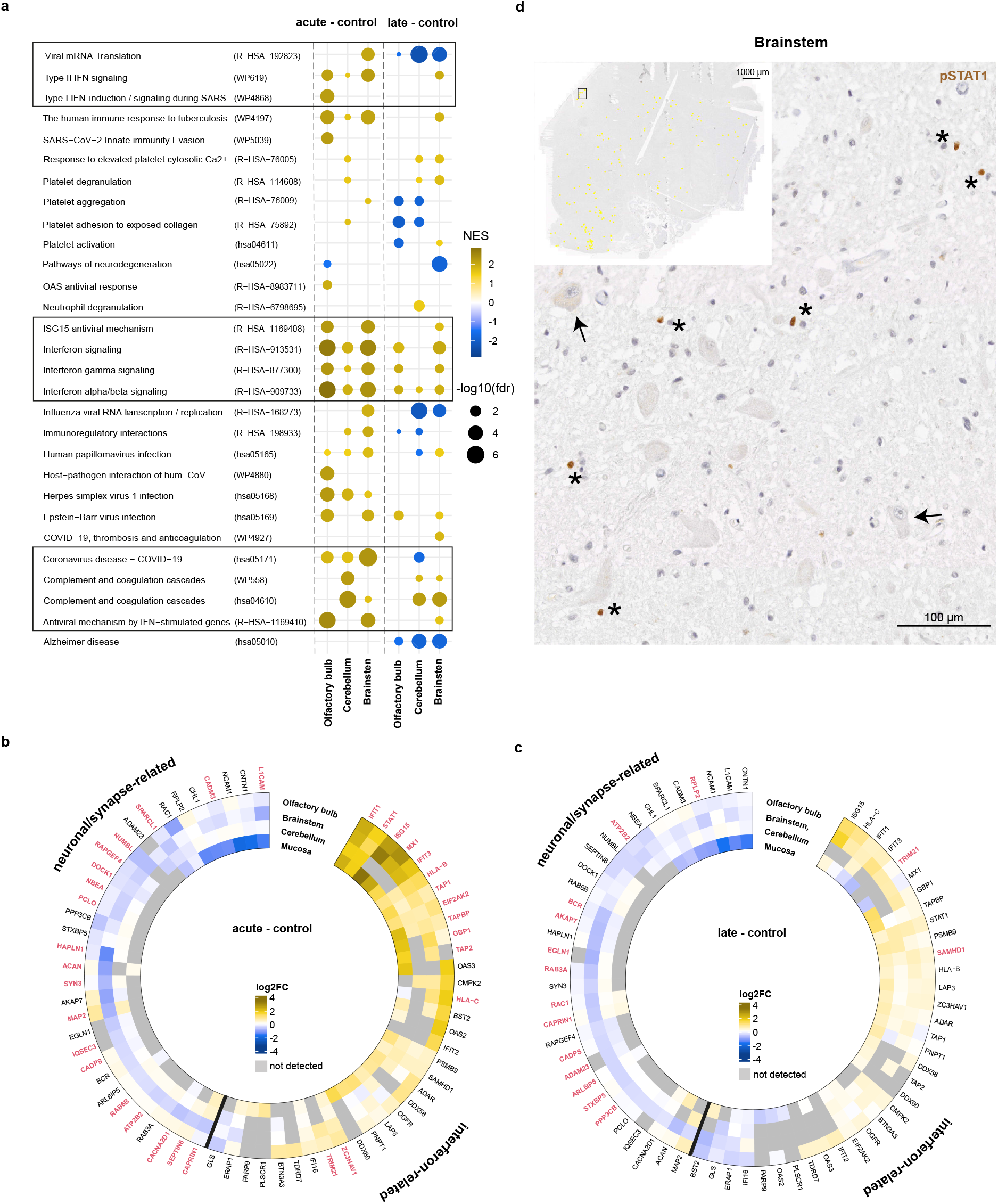
Proteomic landscape and evaluation of COVID-19-enriched pathways. Gene Set Enrichment Analysis (GSEA) showing enriched COVID-19-related biological pathways in the different CNS compartments in acute and late COVID-19 compared to controls (FDR ≤ 0.1; olfactory bulb (n = 18), cerebellum (n = 21), brainstem (n = 22), and olfactory mucosa (n = 21)). The pathways discussed in more detail in the manuscript are highlighted by black boxes (**a**). The circular heatmaps present two subsets of dysregulated proteins (interferon-related and neuronal/synapse-related) in all four anatomical sites (olfactory bulb, brainstem, cerebellum, and olfactory mucosa in acute (**b**) and late (**c**) COVID-19. Significantly regulated genes (*p* < 0.05) in the brainstem are written in red. The data was obtained from linear modeling using the R package LIMMA^145^ (see methods section). Immunohistochemistry of phosphorylated STAT1 (pSTAT1, in brown, asterisk) in the brainstem (scale bar: 100 μm) in an acute COVID-19 case (patient D10). The insert shows an overview of the medulla oblongata (scale bar: 1000 µm) highlighting all pSTAT1 positive nuclei (yellow dots) and the region that was magnified (black square). Two exemplary neurons of the cranial nerve nucleus are marked with black arrows (**d**).

### Interferon related proteins are upregulated in the CNS during systemic infection in acute COVID-19

To validate pathway enrichment analysis and further analyze the differential expression of single proteins strongly associated with *‘*Interferon signaling’, we independently generated a list of interferon (IFN) related proteins, using the INTERFEROME v2.01 database^21^ (**Supplementary Table 2**).

We identified well-known IFN pathway related proteins, such as STAT1, MX1, ISG15, OAS3, DDX60, TRIM21, and TAP1, regulated mostly in the acute phase of the disease in different anatomical regions (**Figure 1 b**). Many of these IFN regulated genes (IRGs) are known to play an important role during the initial stages of viral infections. STAT1 - a pivotal regulator of the transcriptional response to IFNs and other cytokines - was found to be equally upregulated at significance level *a* = 0.01 (moderated t-statistics^22^) in all four tissues in the acute phase, which marks a broad reaction of the brain parenchyma independently of a direct viral infection^23^. Other well-known IFN-related proteins, such as IFN-stimulated gene 15 (ISG15), Interferon induced protein with tetratricopeptide repeats (IFIT1), myxovirus resistance protein A (MX1) - a protein with broad antiviral activity and potential biomarker for viral infections^24,25^ as well as IFN-induced dsRNA-dependent serine/threonine-protein kinase (EIF2AK2) - known to directly inhibit viral replication in a wide spectrum of virus families^26–28^ - showed similar upregulation. During the late disease phase, however, the regulation of IFN-induced proteins was less pronounced (**Figure 1 c**). In line with the proteomic data, we found active STAT1 (phospho-STAT1; pSTAT1) in the brainstem in acute cases by immunohistochemistry - mainly in cells with glial morphology (**Figure 1 d**). In addition, we screened the list of differential expressed proteins for neuronal/synapse-related proteins (using SynGO^29^) to investigate possible alterations on neuronal function due to strong interferon responses as described in mouse models where interferon leads to synapse loss^30^. In our proteomic dataset, neuronal/synapse-related proteins were significantly downregulated in the brainstem in acute COVID-19, e.g. L1CAM, SYN3, HAPLN1, or MAP2 **(Figure 1 b)**. Although these proteins showed less regulation in late COVID-19 compared to controls (**Figure 1 c**), we detected alterations even in the late phase with neuronal/synapse-related proteins such as ATP2B2, RAC1, CADPS, or ADAM23 being significantly downregulated in the brainstem.

### COVID-19 induced changes on single-cell level reflect a bystander reaction of the vascular unit, glial cells and neurons during systemic infection in the acute disease phase

To decipher the contribution of different cell types to the reactive changes, we investigated the differential gene expression of IRGs by snRNA-Seq in the brainstem. Our analysis revealed different subsets of neurons and glial cells, as well as macrophages, lymphocytes and cells of the vasculature and leptomeninges (**Figure 2 a**). From the above-mentioned upregulated proteins in the brainstem, we found a corresponding differential expression of gene levels during acute COVID-19 compared to controls in the vasculature, macrophages/microglia and neurons. *STAT1* was significantly upregulated in venous endothelial cells (*p*=1.29&10^−18^), *EIF2AK2* in macrophages (*p*=6.72&10^−8^) and microglia (*p*=0.001), *MX1* in macrophages (*p*=0.03), and *ISG15* in macrophages (*p*=0.001), venous endothelial cells (*p*=6.78&10^−14^) and glutamatergic neurons (*p*=0.007; **Supplementary Table 3**).

**Figure 2:**
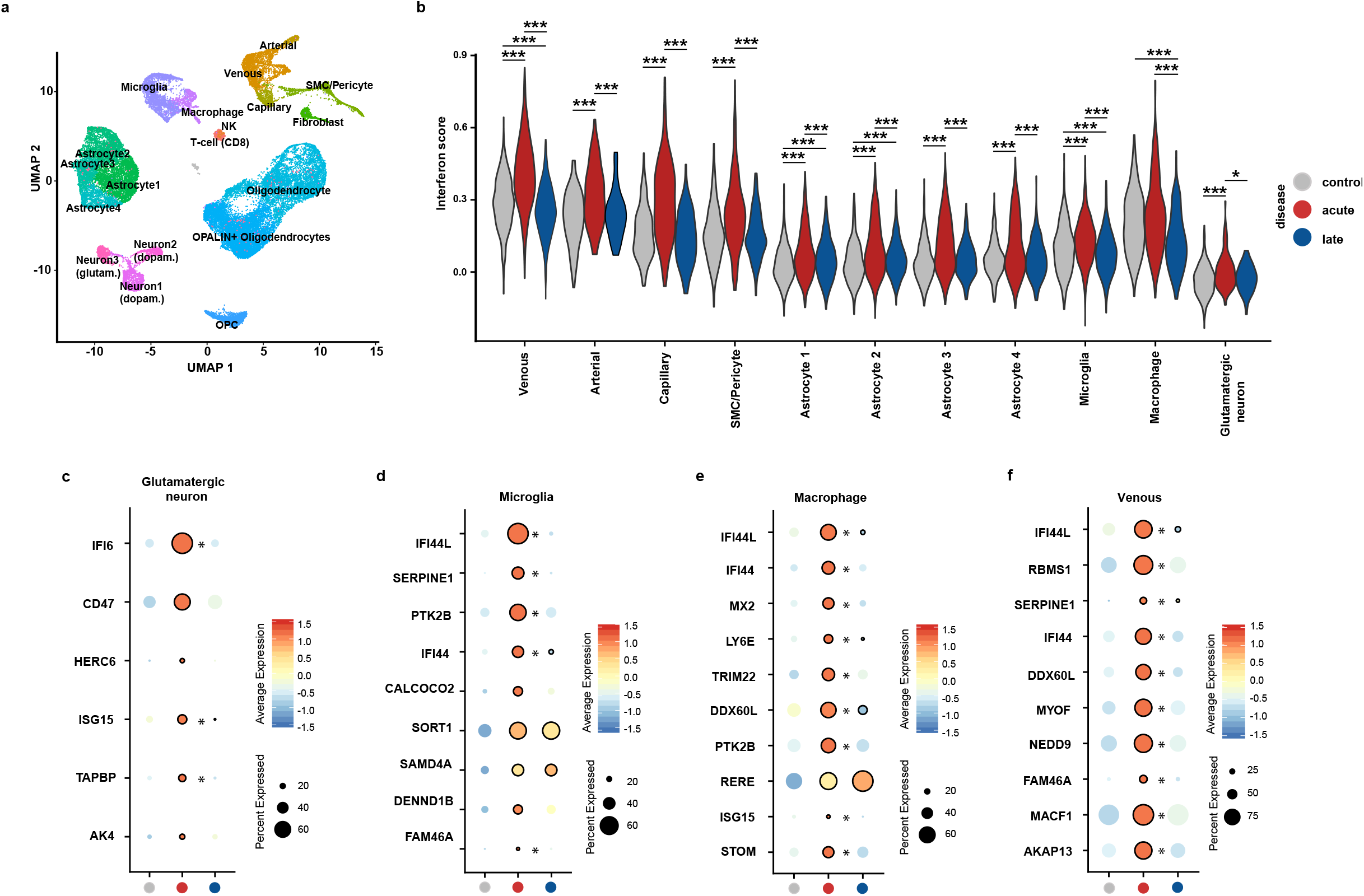
Differential regulation of IFN response in the brainstem. UMAP (Uniform Manifold Approximation and Projection) embedding of the snRNA-seq dataset showing the cellular composition of the brainstem (medulla oblongata, n = 12; **a**). Violin plot comparing the expression of IFN score in acute (n = 4) and late (n = 4) COVID-19 and in controls (n = 4) across different cell populations within the brainstem (**b**). Significance was calculated using the Mann-Whitney U test, the p-values were corrected with FDR. Asterisks indicate level of statistical significance: \*\*\**p* < 0.001, \*\**p* < 0.01, and \**p* < 0.05 (for the Wilcoxon effect size, please refer to **Supplementary table 7**). Dot plots showing the top ten differentially expressed IFN-I-related genes in selected cell types (MAST; FDR adjusted *p*-value < 0.05; **c-f**). Expression levels are color-coded; the percentage of cells expressing the respective gene is size coded. Significant differences are depicted by black circle outlines if a condition is significantly regulated compared to controls and an asterisk if a significant difference is noted between acute and late COVID-19.

To expand the number of integrated candidates involved in the IFN response, we calculated a score of an interferome gene set, which we generated with the INTERFEROME v2.01 database^21^ (see Method section). Using this approach, particularly microglia/macrophages, the vasculature, and astrocytes demonstrated a significant IFN gene set enrichment (*p*<0.05; with moderate to large effect sizes; **Figure 2 b, Supplementary Figure 2 a**) which is in line with other viral infections^31^ and has also been reported in other organs and cell types obtained from deceased COVID-19 patients^32,33^. Interestingly, the glutamatergic neurons showed the strongest alteration among the neuronal cell types and a significant IFN gene set enrichment in the acute phase (**Figure 2 b, Supplementary Figure 2 a**). In the absence of evidence for a direct viral CNS infection and overt peripheral immune cell invasion histologically (deposited at Zenodo, doi: 10.5281/zenodo.7524299) this response is most likely not directly viral-mediated. Notably, bystander inflammatory responses of the CNS to systemic inflammation are also described in septic encephalopathy^34^.

The snRNA-seq dataset showed different interferon related gene patterns in different cell types reflecting a cell type-specific IFN response in the CNS. Intriguingly, the glutamatergic neurons upregulated *CD47* (**Figure 2 c**). The encoded CD47 protein has been described as a “don’t eat me signal”^35^, which is associated with a neuronal-microglial interaction that prevents inappropriate synaptic stripping by microglia via the cognate receptor CD172a^36,37^, thus pointing to neuroprotective mechanisms^38,39^. Moreover, this neuronal cell cluster showed significantly upregulated antiviral cytokines such as *ISG15*, as well as *B2M* (**Supplementary Table 3**) which is not only a component of the MHC class I receptor complex upon viral infection, but also influences neuronal activity^40^.

Microglia, macrophages and endothelial cells (**Figure 2 d-f**) significantly upregulated *IFI44L* and *IFI44*, which can act as negative feedback regulators of host antiviral responses and lead to more viral replication and reduced IFN responses in cell culture experiments^41,42^. Besides the Interferon signature, microglial cells showed a reactive phenotype in the acute phase by downregulating homeostatic markers (*P2YR12* and *CX3CR1*)^43,44^ and upregulated metabolic markers known to be associated with activation such as *APOE*^45^ (**Supplementary Figure 2 b**).

Taken together, our snRNA-seq results demonstrated that within the brainstem, glutamatergic neurons and endothelial cells respond to systemic inflammation in addition to reactive microglia, irrespectively of active SARS-CoV-2 replication. Inflammatory-reactive transcriptomic signatures remained to some extent also in the late phase, characterized by significantly upregulated single IRGs in a small fraction of cells (**Figure 2 c-f**) similar to less pronounced interferon related changes on protein level (**Figure 1 a and d**) in the brainstem.

### Different spatial distribution of interferon related cellular signatures during COVID-19

To evaluate the spatial component of the reaction of CNS cells to the systemic infection, we integrated snRNA-seq data and spatial transcriptomics of the brainstem (**Figure 3 a**). The latter was manually aligned according to the histological image and annotated with the corresponding morphological structures (**Figure 3 b**). The immune-reactive neuron signatures (glutamatergic neurons) from the snRNA-seq dataset were mainly found within the cranial nerve nuclei, particularly within the dorsal nucleus of the vagal nerve as well as the ambiguous nucleus. In contrast, the dopaminergic neurons signatures were found within all brainstem nuclei including the inferior olive. Reactive microglia signatures were not restricted to areas of immune-reactive neurons and found diffusely throughout the white matter (**Figure 3 c**). An unsupervised automated clustering approach revealed phenotypically similar cell clusters to the manually annotated regions, including a cluster of ventral brainstem nuclei (cluster 8) as well as a cluster overlapping with the snRNA-seq signature of dopaminergic neurons (cluster 1), microglia enriched (cluster 7) and white matter fiber tracts (cluster 6; **Supplementary Figure 2 c and d**). Notably, the top 5-20 differentially expressed genes of the microglia enriched cluster (cluster 7) included genes previously described in the context of a reactive microglia phenotype (*APOD, SPP1*, **Supplementary Table 10**).

**Figure 3:**
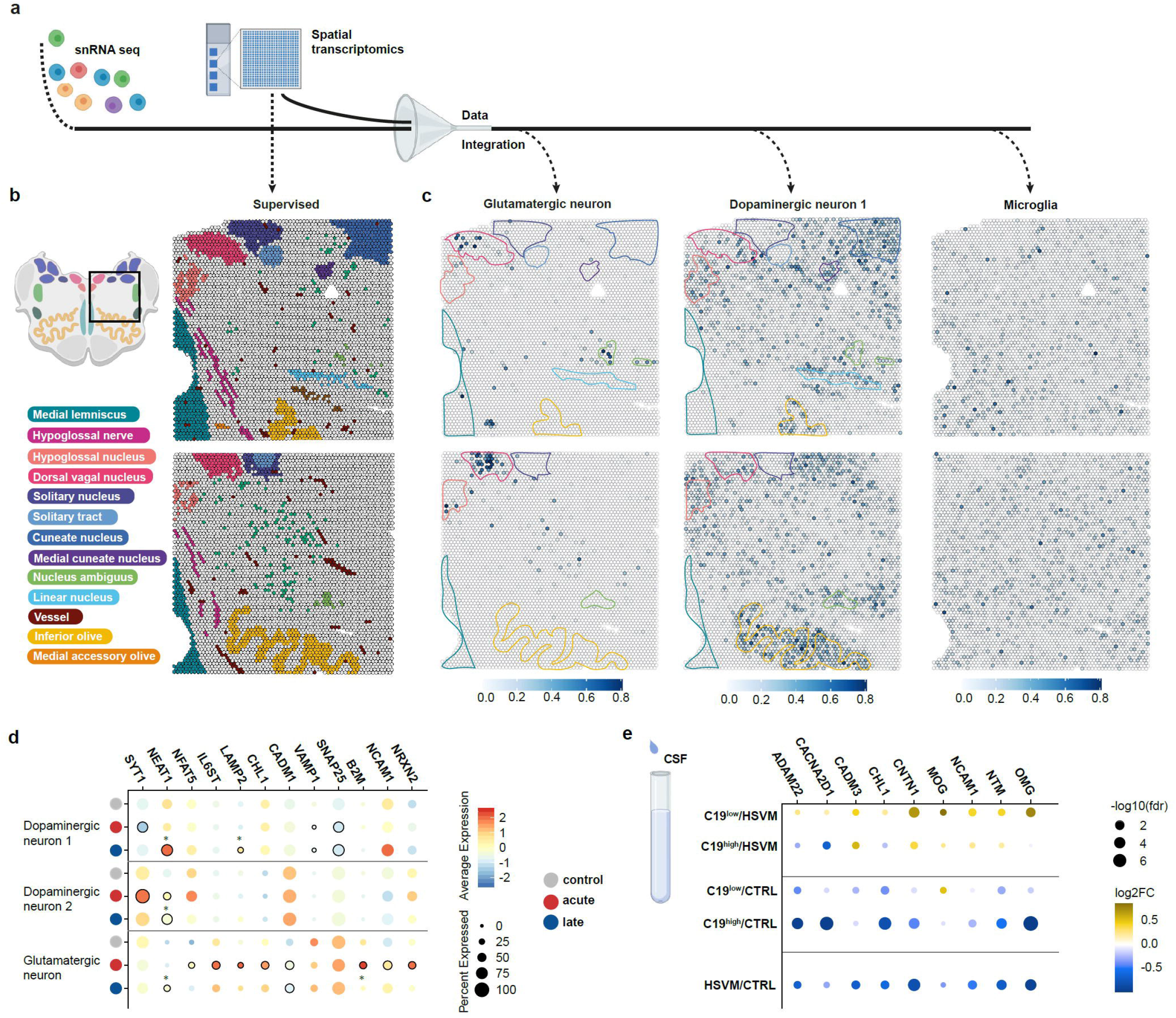
Deregulation of proteins and genes involved in synaptic processes during COVID-19. Schematic representation of the workflow (**a**) with deconvolution of spatial transcriptomics with supervised annotation and color coding of the anatomical regions of the brainstem (medulla oblongata) based on conventional histology and cytoarchitecture used to evaluate the data-driven clustering results (**b**, upper panel: acute (patient D10), lower panel: late (patient D19)) and additional integration of snRNA sequencing results with spot plots depicting the expression patterns for selected neuronal marker genes used to identify dopaminergic and glutamatergic neuronal subsets (in selected schematic color coded anatomical sites) and microglia (**c**). Expression of selected neuronal/synapse-related genes in different neuronal subclusters (glutamatergic, dopaminergic, acute (n = 4) and late (n = 4) COVID-19 and controls (n = 4); **d**). Expression levels are color coded; the percentage of cells expressing the respective gene is size coded. Significant differences are depicted by black circle outlines if a condition is significantly regulated compared to controls and an asterisk if a significant difference is noted between acute and late COVID-19 (MAST; FDR adjusted p-value < 0.05). Dot plot showing the expression of selected myelin- and neuronal/synapse-related proteins in the cerebrospinal fluid (CSF) of living patients with COVID-19 (n = 38) with either high (C19high) or low (C19low) serum levels of procalcitonin (PCT) compared to non-inflammatory controls (CTRL, n = 28) as well as patients with herpes simplex virus meningitis (HSVM, n = 10; **e**).

### COVID-19 induces impairment of synaptic processes in the brainstem

As we found neurons in the cranial nerve nuclei reacting to the systemic inflammation, we explored our snRNA sequencing and proteomic datasets for changes in expression of synaptic proteins. Among a list of proteins identified using SynGO^29^, we observed a downregulation in protein expression associated with synaptic processes, such as SYN3 or L1CAM, both in the acute and late disease phase (**Figure 1 b and c**). The affected proteins in the proteomic dataset were of pre- and postsynaptic origin and were mainly involved in synaptic organization and presynaptic vesicle cycle (**Supplementary Figure 3 a and b**), suggesting an impairment of synaptic processes in the brainstem in the acute disease phase and during the entire disease duration.

On transcriptome level, the different neuronal cell clusters showed a distinct regulation of genes associated with synapse biology. Specifically, synaptosomal-associated protein of 25kDa (SNAP25), vesicle-associated membrane protein 1 (*VAMP1*) and Synaptotagmin-1 (*SYT1*) - components of the SNARE complex - were significantly downregulated in dopaminergic neurons (**Figure 3 d**). The SNARE complex is involved in synaptic vesicle exocytosis, a fundamental step in neurotransmitter release^46^. Our findings are in line with previously published results by Yang *et al*. describing the downregulation of synaptic genes that mediate neurotransmission in cortical neurons of post-mortem tissue in COVID-19 patients^8^. These alterations were found in both, the acute and late phase of the disease (**Figure 3 d**). Furthermore, dopaminergic neurons (neuron cluster 1 and 2) showed significant changes compared to the control group in *NEAT1* expression, a long non-coding RNA, described to modulate human neuronal activity^47^ as well as *SYT1*, a critical mediator of neurotransmitter release^48^.

In contrast, glutamatergic neurons showed significantly higher expression of Neurexin 2 (*NRXN2*), cell adhesion molecule 1 (*CADM1)*, Neural Cell Adhesion Molecule 1 (*NCAM1*), and Cell Adhesion Molecule L1 Like (*CHL1*) in the acute disease phase (**Figure 3 d**). These adhesion molecules are involved in synaptic organization and signaling. For instance, Neurexins serve as presynaptic receptors for several extracellular binding partners, indicating their essential role in presynaptic organization^49,50^.

To validate our findings regarding synapse and myelin related proteogenomic alterations, we analyzed the proteomic profile of CSF in an independent COVID-19 cohort. In line with our post-mortem analysis, synaptic proteins interacting with the SNARE complex, such as CHL1^51^, were almost non-detectable in the CSF of patients with non-lethal COVID-19 (with and without concomitant bacterial superinfection) (**Figure 3 e;** −log10 (FDR) >4). Moreover, detection of proteins such as Oligodendrocyte-myelin glycoprotein (OMG) and Contactin-1 (CNTN1) were reduced in the CSF. OMG is highly expressed on the surface of oligodendrocytes and neurons, regulating synaptic plasticity^52^. CNTN1 is also expressed on neurons and has been reported to be involved in myelin formation^53^. Therefore, reduced CNTN1 in the CSF could be a sign for altered myelin homeostasis in the CNS during COVID-19, in line with alterations of myelin formation in the snRNA dataset where we detected a significant decrease in frequencies of (remyelinating) *OPALIN+* oligodendrocytes in acute COVID-19 (**Supplementary Figure 3 c; Supplementary Table 8**).

### Response of the perivascular unit in the brainstem during COVID-19 disease course

Endothelial cells and macrophages showed the strongest interferon related signature (**Figure 2 b**). Therefore, we focused on the perivascular space as an immune-regulatory unit in more detail. Endothelial cells in the brainstem upregulated genes encoding for integrins (*ITGA1, ITGA5, ITGA6, ITGA9, ITGAV*) and adhesion molecules (*ICAM-1, ICAM-2, SELE*) (**Figure 4 a, Supplementary Table 3**). This so-called ‘type II activation’ is necessary for recruiting immune cells from the periphery^54,55^ and more pronounced in the acute than in the late disease phase. Accordingly, arterial and capillary endothelial cells obtained from donors with acute COVID-19 upregulated *ANGPT2* (Angiopoietin-2), encoding a protein involved in endothelial cell activation, sensitization to circulating cytokines, and promotion of angiogenesis^56–58^. Peripherally increased Angiopoietin-2 is a surrogate marker of endothelial activation and indicator of worse outcome in COVID-19^59^. Moreover, *TIE1*, encoding a receptor for Angiopoietins 1 and 2 and reported to regulate the expression of adhesion molecules^60,61^, was upregulated in capillary endothelial cells. This cluster also demonstrated a strong upregulation of *MCAM* (melanoma cell adhesion molecule; CD146), which encodes a molecule important for T cell recruitment to the CNS as well as a cell surface co-receptor for well-known endothelial growth factors such as VEGFA, VEGFC, PDGFB, and FGF4^62–66^ (**Figure 4 a**).

**Figure 4:**
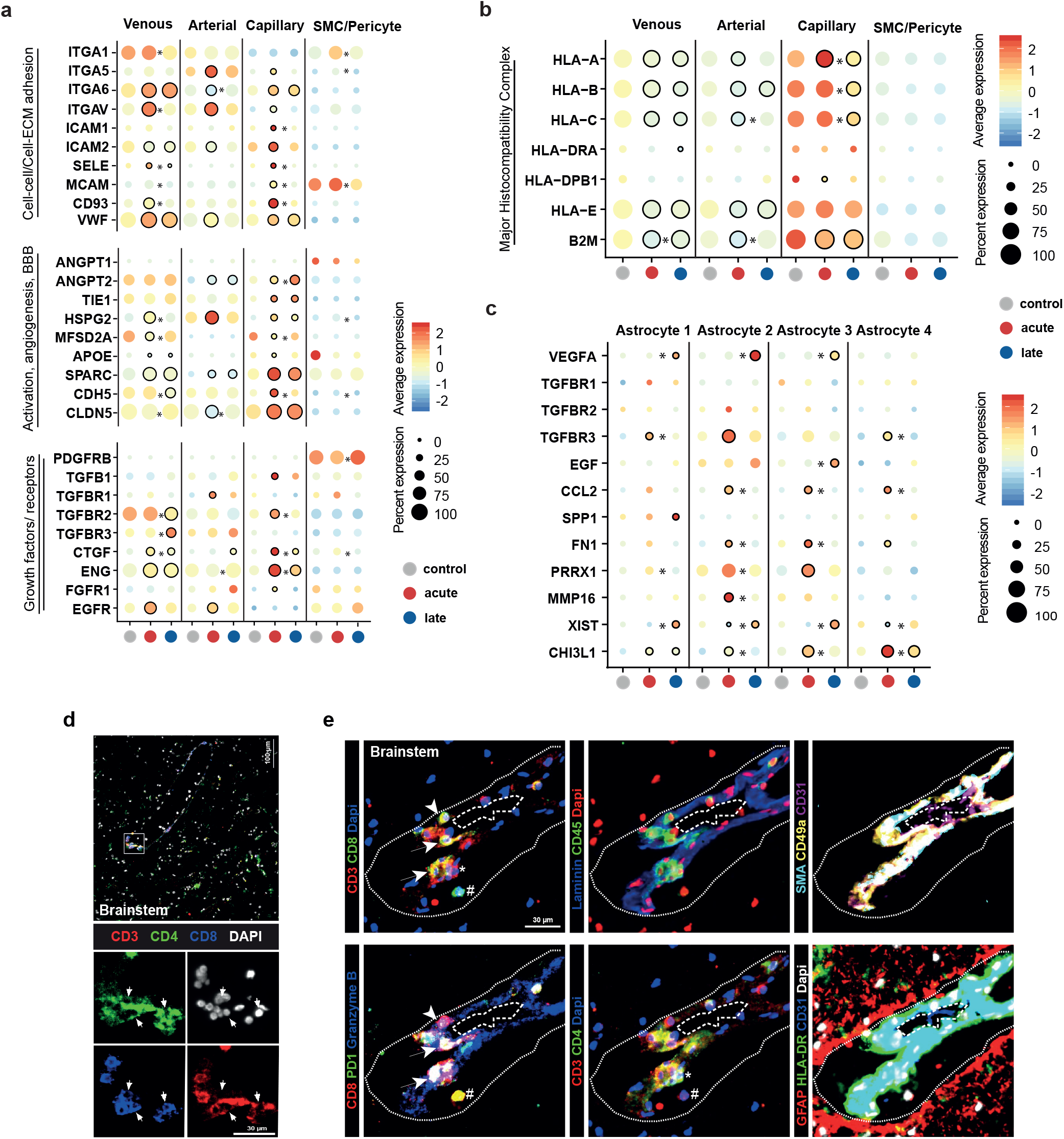
Perivascular unit in the brainstem shows inflammatory changes during COVID-19 disease course. Dotplot visualization of gene expression for various genes of interest in different endothelial cell subsets (venous, capillary, arterial) and SMC/Pericytes (**a, b**) and astrocytes (**c**) during COVID-19 (acute (n = 4) and late (n = 4) COVID-19 and controls (n = 4)). Expression levels are color coded; the percentage of cells expressing the respective gene is size coded. Significance is depicted by black circle outlines if a condition is significantly regulated compared to control and a star (*) if a significant difference is noted between acute and late COVID-19 (MAST; FDR adjusted p-value < 0.05). (BBB: blood brain barrier; ECM: extracellular matrix). An overlay (marked as square, upper panel) and single channels (lower panel) of multiplex fluorescence images (MELC) obtained from one single section and one field of view of the brainstem (patient D16 late) highlights double positive T cells expressing CD4+ and CD8+ (arrows, (**d**)). Overlays of MELC images obtained from one single section and one field of view of the brainstem (patient D18 late). CD8+ T cells heterogeneously expressing PD-1 and Granzyme B are indicated by arrows and an arrowhead. A hashtag marks one CD8+ T cell only expressing PD-1 and no Granzyme B. One CD4+ T cell is marked by an asterisk. The dashed line reflects the CD31+ endothelium, the dotted line represents the Virchow-Robin space (**e**).

In addition to the strong type I IFN signature described above, capillary endothelial cells in the brainstem of patients with an acute disease course showed a significant increase in the expression of transforming growth factor-β (TGF-β) coding gene *TGFB1*. We also found an upregulation of the corresponding receptors (*TGFBR1, TGFBR2, TGFBR3*) in venous and arterial endothelial cells as well as in subpopulations of astrocytes (**Figure 4 a and c**). Besides, we observed a downregulation of *MFSD2*A and *CLDN5* in endothelial cells in the acute phase and an upregulation of *SPARC* in both, the acute and late phase as well as a downregulation of *APOE* in SMC/pericytes (**Figure 4 a**). These alterations suggested an increased leakiness of the blood brain barrier (BBB)^67–75^, which has also been hypothesized by others in severe COVID-19^76^. In addition, recent CSF studies support this dysfunction showing a leaky BCB (blood cerebrospinal fluid barrier) with elevated proteins and cells coming from the periphery^77^. The constituents of the major histocompatibility complex (MHC) were significantly downregulated in both disease phases (**Figure 4 b**). Finally, Endoglin (*ENG*), a co-receptor of TGFβ^78–80^, and *CTGF* (connective tissue growth factor) were found upregulated in venous and capillary endothelial cells, as well as in SMC/pericytes (**Figure 4 a**). Beside their known role in tissue fibrosis^81,82^, CTGF and TGFβ have been described to play a role in activation of astrocytes and microglial cells^83^.

In fact, the perivascular space is the main homeostatic niche for tissue-resident memory T cells^84^. The alterations of the vascular unit pointed to an increased immune cell recruitment from the periphery with upregulation of integrins but with reduced local reactivation in the perivascular space due to downregulation of MHC molecules (**Figure 4 b**). An integrated UMAP of the snRNA-seq data showed that cerebral, pulmonary and mucosal T cells clusters contain NK cells, CD4+ and CD8+ T cells (**Supplementary Figure 4 a**). However, there were only few commonly up- or downregulated genes throughout these organs (**Supplementary Figure 4 b**) with upregulation of activation markers such as *CCND3* or *IFI44L*. As expected, we only found very few T cells in our brainstem snRNA-seq dataset compared to mucosa and lung limiting the gene expression analysis of activation and cytotoxic markers (**Supplementary Figure 4 c and d**), which is why we additionally performed multiplex histology in a stain and bleach approach^85^ to further characterize these cells (**Figure 4 d and e**). Also in histology we only found single CD3+ cells, mainly in the perivascular space in the brainstem. We found CD4+ T cells but we also detected double positive (DP) CD4+ and CD8+ T cells in the perivascular space (**Figure 4 d**). DP T cells are a subpopulation known to accumulate in lung tissue of COVID-19 patients^20^ and various other disorders^86^ producing pro-inflammatory cytokines and displaying characteristics of memory CD8+ T lymphocytes^86^. Similar to blood derived T cells in severe COVID-19^85^, we identified T cells expressing granzyme B and PD-1, suggesting a still ongoing local (unconventional) activation within the perivascular space even in the late disease phase (**Figure 4 e**).

## Discussion

Although the brain is not the primary target of SARS-CoV-2^9^, the strong systemic inflammatory response in severe COVID-19 can be detrimental to CNS function and integrity (summarized in **Figure 5**). SARS-CoV-2 infection is associated with neurocognitive and neurological impairment in a subset of patients in the absence of severe morphological changes and viral replication in neurons, glial or other CNS cells^7^. Albeit low level of viral RNA was detected by ultrasensitive methods such as RT-qPCR in 4 out of 21 brainstem samples (**Supplementary Table 1**), viral protein (immunohistochemistry; deposited at Zenodo, doi:10.5281/zenodo.7524299) or viral transcripts (snRNA-Seq) were not found in the CNS (**Supplementary Figure 5**), which is in line with previous results by Yang *et al*.^8^. Additionally, not the viral RNA load but disease duration significantly shaped the proteomic profile of the CNS (**Supplementary Figure 1**) suggesting a stronger impact of peripheral inflammatory changes during the acute phase of systemic infection.

**Figure 5:**
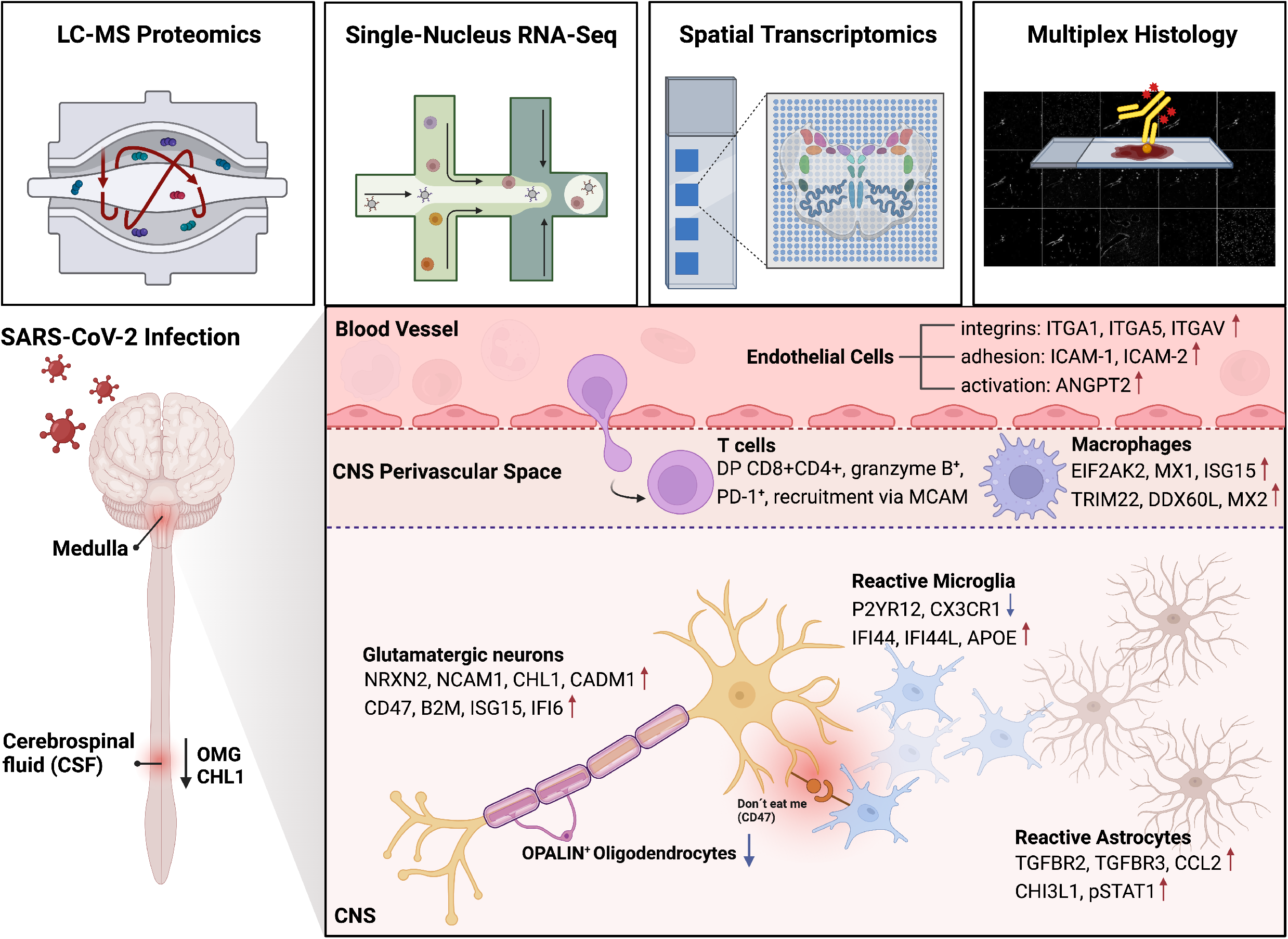
Study design and multi-omic approach. The image summarized the study design, methods used and important cell-specific results from the multi-omic analysis of the medulla oblongata from patients that died of COVID-19, showing strong activation of endothelial cells, increased permeability of the blood brain barrier (BBB), altered T cell and microglial activation, and neuroprotective mechanisms such as neuronal upregulation of CD47. Important up- and downregulated genes and/or proteins are shown for certain cell types in the different CNS compartments.

Our results demonstrate a proinflammatory CNS microenvironment during systemic COVID-19 infection with significant upregulation of IFN-stimulated genes and proteins as well as significant downregulation of genes and expressed proteins associated with synaptic organization and myelination. However, we did not see a marked increase in T cell numbers or additional microglial populations as previous studies (**Supplementary Figure 3 c**)^8,14,87,88^ probably due to our control group with also multi-organ failure, superinfection and to some degree neurodegenerative diseases similar to the COVID-19 groups (**Supplementary Table 1**). The described IFN response in our study is probably not SARS-CoV-2-specific and not only seen in severe COVID-19 as similar alterations of neurons and oligodendrocytes were previously described in CNS viral infections caused by the West Nile virus (WNV)^31^ and - in the CSF - by Herpes virus (HSV; **Figure 3**)^10^. Interestingly, during COVID-19 we found a tendency to reduced glutamatergic neurons (not significant in our testing) but we found a net reduction of *OPALIN+* oligodendrocytes, a finding previously reported in demyelinating lesions and ‘normal appearing white matter’ in multiple sclerosis (MS)^89^. *OPALIN* is a known marker gene expressed by myelinating oligodendrocytes and induced during oligodendrocyte differentiation^90^. The marked reduction of this cell cluster and reduced CNTN1 in the CSF (**Figure 3 e**) point to an altered myelin homeostasis. *OPALIN+* oligodendrocyte reduction and reduced CNTN1 in the CSF are both also described in demyelinating conditions such as MS^89,91^. However the altered myelin homeostasis in COVID-19 is more subtle and does not lead to overt demyelination detectable by conventional histology. In our cohort, numbers of remyelinating oligodendrocytes (*OPALIN+*) recovered in late COVID-19 pointing to a reversible process. In contrast, neurons do not regenerate and require adaptive protective processes. One possible adaptation additional to *CD47* upregulation could be the reduced expression of L1CAM protein in acute COVID-19, which is described to be an adaptive process of neuronal self-defense in response to neuroinflammation in mice^92^. On the other hand, the described alterations in gene and protein expression associated with synaptic organization and myelination could lead to functional impairment of the reticular formation, which is involved in cardiovascular control, sleep and consciousness^93^.

While there is no robust evidence for a direct infection of endothelial cells by SARS-CoV-2^94–96^, multiple studies suggest capillary damage in various organs^97^ and functional impairment of the microvasculature has been implicated in the pathogenesis of COVID-19 acutely in the pandemic^98–102^. We didn’t find overt signs of neurovascular damage, increased thrombosis, endothelial destruction or bleedings in our brainstem samples. However on a transcriptional level, besides the response to interferon, we find signs of BBB damage. During the acute disease phase, endothelial cells in the brainstem display multiple signs of activation in the absence of SARS-CoV-2 protein or RNA. *TGFB1* is upregulated in endothelial cells in the acute disease phase, which appears to be a major immunopathological mechanism in severe COVID-19^85^. Actually, high serum levels of TGFβ are detected in the first two weeks of SARS-CoV-2 infection and NK cells show a dominant TGFβ response signature, which seems to inhibit NK cell function and early control of the virus^103^. Furthermore, we find a downregulation of MHC class I molecules (e.g. HLA-A, -B, -C, -DRA) in endothelial cells in the brainstem, which may also be explained by the activation of TGFβ signaling^104^ within the vascular unit. These results suggest that suppression of cellular inflammation within the CNS through the vascular unit may prevent its collateral damage^105,106^. However, unconventionally activated T cells are continuously detectable also in the late COVID-19 cases in our cohort. In mice, a recent study described cerebral T cells as inducers for IFN responsive microglia and oligodendrocytes^107^. Perhaps in COVID-19, the cerebral perivascular unit with its CD8+ T cells (together with endothelial cells, macrophages and astrocytes) also induces the described inflammatory response in the brain, explaining the diffuse distributed reactive microglial signatures in our spatial sequencing dataset independent from the ‘interferon-reactive’ neurons located in the cranial nerve nuclei (**Figure 3 b and c**). These neurons could get activated independently and promote resolution of inflammation as described in animals with the “inflammatory reflex”^108^, where peripheral inflammation leads to electrical stimulation of vagal neurons that lead to modulation of inflammation.

Although our results clearly point to a significant upregulation of SARS-CoV-2 induced inflammatory reactions mostly in the acute phase of disease, one limitation of the present study is the sparse clinical information about the patient’s neurological state. Our findings on the protein and RNA level are in line with two previous CNS snRNA sequencing studies^8,87^, which also show upregulation of inflammatory genes in all investigated cell types as well as alterations of excitatory neurons in cortical layers II-IV, which are linked to cognitive function^109^. It is currently discussed if the SARS-CoV-2 pandemic will lead to a second pandemic of neurodegenerative diseases in the next years but further studies are needed to investigate the direct link of viral induced systemic inflammation and its effects on myelination, synapse organization and neurodegeneration and to correlate the clinically observed neurological symptoms and changes in cognition due to SARS-CoV-2 infection as well as other virus induced or autoimmune/autoinflammatory disorders. Along this line, certain “interferonopathies’’ such as Aicardi-Goutières syndrome and SLE are associated with neurological symptoms and cognitive impairment^110–112^ and studies in mice suggest a detrimental collateral damage of the CNS in the context of a systemic type I Interferon response in certain viral infections^113–116^. In mice, also chronic presence of IFN-I in the brain alone could negatively affect cognitive function, mediated via modulation of microglial activity^117^.

Briefly, this indicates that systemic inflammation induced by type I Interferons perturbates CNS homeostasis on a cellular and molecular level, potentially explaining neurocognitive disturbances in patients with severe COVID-19. This could also support the idea of treating patients with severe systemic infection-induced encephalopathy with immune modulators such as intravenous immunoglobulins, which showed a positive effect in small case series^118,119^ as they are known to inhibit with interferons^120^.

## Material and Methods

### Post mortem study design

We analyzed autopsy tissue samples from n = 23 male and n = 7 female individuals. The detailed clinical information as well as general confounding factors and comorbidities relevant for neuroinflammation are given in **Supplementary Table 1**. Patients with COVID-19 (n = 21) were divided into two groups, namely ‘acute’ (n = 10; median age: 77.5 years, age range: 63 - 91 years) and ‘late’ (n = 11, median age: 78 years, age range: 56 - 92 years) depending on whether death occurred within 14 days from the beginning of symptoms or more than 14 days, respectively. All procedures performed in this study were in accordance with the ethical standards of the respective institutional research committees and with the 1964 Helsinki declaration and its later amendments or comparable ethical standards. The study was approved by the local ethics committees at Charité (approval numbers: EA1/144/13, EA2/066/20, EA1/075/19) and the Charité-BIH COVID-19 research board. Informed consent for autopsy was obtained for all individuals. Randomization was not performed as all individuals with detectable SARS-CoV-2 RNA by RT-PCR were included in the COVID-19 group and controls were defined as individuals negative for SARS-CoV-2 by RT-PCR. At least two independent board-certified neuropathologists histologically assessed all tissue samples.

### CSF sample collection

In a retrospective study, we collected CSF samples from patients with a PCR-proven SARS-CoV-2 infection (nasopharyngeal swab testing) who underwent lumbar puncture to rule out CNS pathologies such as autoimmune encephalitis or meningitis, involving a total of n = 38 COVID-19 patients, as well as patients with herpes simplex virus (HSV) encephalitis (n = 10) and non-inflammatory controls (n = 28). Cohort characterization is provided in **Supplementary Table 5**. We divided the COVID-19 cohort into two subgroups, namely ‘PCTlow’ and ‘PCThigh’ depending on serum procalcitonin (PCT) levels. Low PCT levels (<0.5 µg/L) are a surrogate for non-septic conditions, whereas elevated PCT levels (>1.0 µg/L) indicate a high likelihood of systemic bacterial infection. This part of the study was approved by the local ethics committees at Charité (EA1/76/20). And these samples are also included in a separate CSF study^10^.

### Single-nucleus RNA Sequencing (snRNA-seq)

For snRNA-seq, we used fresh frozen samples of brainstem (medulla oblongata n = 8; acute n = 4, late n = 4) and olfactory mucosa (n = 6; acute n = 3, late n = 3) from deceased COVID-19 patients and controls (medulla oblongata n = 4; olfactory mucosa n = 3. Additional a lung dataset was included from our previous publication^121^. Brainstem and mucosal nuclei were isolated and prepared for snRNA-seq as previously described^122^. Briefly, 60-70 μm thick tissue sections were homogenized with glass tissue douncers in citric acid buffer (250 mM sucrose, 25 mM citric acid, 1 μg/mL Hoechst 33342). The homogenate was successively passed through a 100 μm and a 35 μm cell strainer to remove large debris. This was then centrifuged for 5 minutes at 500 x g at 4°C. The nuclei were then washed in 1 ml of citric acid buffer. To remove residual debris, the nuclei were passed through a density centrifugation gradient by adding an equal volume of S88 citric acid buffer (882 mM sucrose, 25 mM citric acid) at the bottom of the tube before being centrifuged for 10 minutes at 1000 x g at 4°C. The supernatant was carefully removed, and the nuclei were resuspended in cold resuspension buffer (25 mM KCl, 3 mM MgCl2, 50 mM Tris-buffer, 0.4 U/μL RNaseIn Takara 2313A, 1mM DTT, 0.4 U/μL SuperaseIn Thermo Fisher Scientific AM2694, 1 μg/mL Hoechst 33342). The nuclei were counted and then immediately loaded into the 10x Chromium Controller using the 10x Genomics Single Cell 3’ Library Kit v3.1 (10x Genomics; PN 1000223; PN 1000157; PN 1000213; PN 1000122). Subsequent steps were performed according to the manufacturer’s instructions with minor modifications. Specifically, the 85°C incubation step during the reverse transcription step was extended to 10 minutes and an additional 2 cycles were added to the cDNA amplification. Afterwards, the libraries were pooled and sequenced on the NovaSeq 6000 Sequencing System (Illumina, paired-end, single-indexing).

After pre-processing and quality control steps, snRNA-seq yielded a total of 52,421 single-nucleus transcriptomes in the olfactory mucosa and 36,780 single-nucleus transcriptomes in the medulla oblongata. Through unsupervised clustering and the use of known marker genes, 27 distinct clusters in the olfactory mucosa and 20 clusters in the medulla could be identified^121,123–127^. A cluster of cells were left undefined that did not express known marker genes of brain cell types. Microglia/macrophage cluster 1 shows less expression of *PTPRC (CD45), CD47, ITGAM (CD11b), CD14* and *CD68* compared to macrophage/microglia cluster 2 and more expression of *P2YR12* and *TMEM119*, so we assume that cluster 1 contains more ‘microglia’ and cluster 2 more ‘macrophages’^37^ (**Supplementary Figure 7c and h**).

In the medulla dataset endothelial cell cluster 1 shares features of venous endothelial cells, cluster 2 of arterial endothelial cells and endothelial cluster 3 of capillary endothelial cells^128^ and were named accordingly. The neuronal clusters show no exclusive neurotransmitter profiles, however neuron cluster 1 and 2 show enriched expression of genes related to dopaminergic neurons and neuron cluster 3 of glutamatergic neurons^129–136^ (**Supplementary Figure 7 c and i**).

### Interferon (IFN) score

To generate the IFN score, all differentially upregulated genes in COVID-19 acute compared to the control (logFC > 0.25) were obtained and parsed through the INTERFEROME database to identify IRGs^29^. This IFN gene list was then used to calculate a score using the AddModuleScore function in Seurat^137^. This generates an average expression for the IFN program that is subtracted by the aggregate expression of control genes.

### Proteomics

75 µl of tissue lysis buffer (Covaris 520284) were added to approximately 5 mg of different tissue types in AFA-TUBE TPX 8-strips (Covaris 520292; (olfactory mucosa n= 22, olfactory bulb n = 18, brainstem n = 24, cerebellum n = 21; **Supplementary Table 1**). Proteins were extracted on a Covaris LE220Rsc (3 cycles of 375 PIP, 25% DF, 50CPB, 20 repeats, 10 s pulse duration, 10 s delay, 12°C, 3 mm dither @ 20 mm/s). Three samples (n=1 mucosa D20 and n=2 brainstem D12 and D8) were excluded from the analysis due to low quality (**Supplementary Table 1**). Strips were shortly centrifuged and proteins quantified (Pierce Protein Assay Kit, 23225). 25 µg of protein was transferred to a 96 well plate and filled to 50 µl with water. The lysates were processed on a Biomek i7 workstation using the SP3 protocol as previously described with single-step reduction and alkylation^138^. Briefly, 16.6 μl reduction and alkylation buffer (40 mM TCEP, 160 mM CAA, 200mM ABC) were added, the samples were incubated for 5 min at 95°C. To bind the proteins, 200 μg paramagnetic beads (1:1 ratio of hydrophilic/hydrophobic beads) were added and proteins precipitated with 50 % ACN. The samples were washed 3 times with EtOH and once with ACN, before reconstitution in 35 μl 100 mM ABC, addition of 2 μl trypsin/LysC (1 μg/ 0,01ug) overnight digestion at 37 °C. The reaction was stopped by adding formic acid up to a final concentration of 0.5 %. Samples were then transferred to a new plate and used for LC-MS/MS analysis without additional conditioning or clean-up.

### Liquid Chromatography-Mass Spectrometry Analysis (LC-MS)

1.25 µg peptides were concentrated for 3 min on a trap column (PepMap C18, 5 mm x 300 μm x 5 μm, 100Ǻ, Thermo Fisher Scientific) with a buffer containing 2:98 (v/v) acetonitrile/water containing 0.1% (v/v) trifluoroacetic acid at a flow rate of 20 μl/min. They were separated by a 250 mm LC column (Acclaim PepMap C18, 2 μm; 100 Å; 75μm, Thermo Fisher Scientific). The mobile phase (A) was 0.1 % (v/v) formic acid in water, and (B) 80 % acetonitrile in 0.1 % (v/v) formic acid. In 128 min total acquisition, time gradient B increased in 90 min to 25 %, and in 20 min to 50 % with a flow rate of 300 nl/ min. For the tissue samples, the Orbitrap worked in centroid mode with 12 m/z DIA spectra (12 m/z precursor isolation windows at 17,500 resolution, AGC target 1e6, maximum inject time 60 ms, 27 NCE) using an overlapping window pattern. Precursor MS spectra (*m*/*z* 400-1000) were analyzed with 35,000 resolution after 60 ms accumulation of ions to a 1e6 target value in centroid mode. Typical mass spectrometric conditions were as follows: spray voltage, 2.1 kV; no sheath and auxiliary gas flow; heated capillary temperature, 275 °C; normalized HCD collision energy 27%. Additionally, the background ions m/z 445.1200 acted as lock mass.

### Computational proteomics

For every tissue, a tissue specific library was constructed using standard settings in library free mode with DIA-NN (version 1.7.16)^139^. The libraries were automatically refined based on the project dataset at 0.01 global q-value (using the “Generate spectral library” option in DIA-NN). The output was filtered at 0.01 false discovery rate (FDR) at the peptide level. For gas phase fractionation (GPF), 6 single 1.25 μg injections of pooled tissue samples were analyzed by online nanoflow liquid chromatography tandem mass spectrometry as described above. For GPF, an overlapping window pattern from narrow mass ranges using window placements (i.e., 395-505, 495-605,595-705, 695-805,795-805,895-905 m/z) was set. Two precursor spectra, a wide spectrum (395-505 m/z at 35,000 resolution) and a narrow spectrum matching the range using an AGC target of 1e6 and a maximum inject time of 60 ms were analyzed every 25 MS/MS with 4m/z precursor isolation windows at resolution of 17,500.

### Proteomics data analysis

DIA_NN normalized peptide intensities were used as input. Each tissue type was processed separately. One mucosa and two brain stem samples were identified as outliers and were excluded. Peptides with excessive missing values (> 40% per group) were not considered in our analysis. The missing values of remaining peptides were imputed group-based using the PCA method^140^. Then they were normalized using *vsn* method^141^. To obtain a quantitative protein data matrix, the log2-intensities of peptides were filtered, only peptides belonging to one protein group were kept, and then summarized into protein log intensity by “maxLFQ” method^142^, implemented in R package iq^143^.

Statistical analysis of proteomics data was carried out in R using publicly available packages. Principal component analysis was based on R package FactoMineR^144^. Linear modelling was based on the R package LIMMA^145^. Following model was applied to each tissue data set (log(p) is log2 transformed expression of a protein): log(p) ∼ 0+Class. The categorical factor Class has three levels: control, early, late; Reference level: late.

For finding regulated features following criteria were applied: Significance level alpha was set to 0.01, which guaranteed Benjamini–Hochberg false discovery rate^146^ below ∼ 30% across all contrasts in the tissue and across all tissues in the contrast. The log fold change filter was applied to guarantee that the measured signal is above the average noise level. As such we have taken median residual standard deviation of linear model: log2(T) = median residual SD of linear modelling (T_bulb_ = 1.37, T_cerebellum_ = 1.23, T_brainstem_ = 1.25, T_mucosa_ = 1.45). Functional GSEA analysis was carried out using the clusterProfiler R package^147^. For selecting the most (de)regulated GO terms we applied filter: 2 ≤ term size ≤ 250.

### Visium Spatial Gene Expression (10 X Genomics)

Spatial transcriptomics were performed following the respective Visium Spatial Protocols (CG000295 Rev D, CG000240 Rev D, CG000160 Rev C, CG000239 Rev E). Fresh frozen samples (n = 7) were cryosectioned and placed on Visium Spatial Gene Expression Slides (10X Genomics, PN-1000184). Tissue sections were then fixed in methanol and H&E stained according to the Visium Spatial Gene Expression User Guide (CG000160 Rev C). For gene expression samples, tissue was permeabilized for 24 minutes, which was selected as the optimal time based on tissue optimization time course experiments. Brightfield histology images were taken right after using a Keyence BZX800. The tissue was captured in brightfield mode with a 10x magnification and stitched with the BZ-X800 Analyzer software. Afterwards, the coverslip was removed by holding the slide horizontal in a beaker containing 800 ml ultrapure water. Permeabilization, reverse Transcription, second Strand Synthesis, Denaturation and cDNA Amplification was performed according to Visium Spatial Gene Expression User Guide CG000239 Rev E.

### Spatial transcriptomics data processing

Fastq output files of the sequenced library, including the spatial barcode, were aligned to the H&E image using Space Ranger. Following downstream analyses, quality control (QC) processing, batch data integration, graph-based clustering, visualizations, and differential gene expression analyses were performed using the Seurat package (v4.0), R, R Studio^137^. We considered only the spots showing the expression of more than 100 genes. In addition, we selected genes that were expressed in at least three sequenced spots. Morphological structures were manually aligned to the H&E image using Loupe Browser v6 (n = 5). To visualize predictions of the cell types using the Seurat package^137^, we integrated the spatial transcriptomic dataset with the snRNA-seq dataset generated from the same brain regions and tissue samples.

### Histological Staining methods

Descriptions of histological staining procedures (pSTAT1, Nucleocapsid and Multiplexhistology) are given in the **Supplementary Material** section.

### Statistics & Reproducibility

No statistical methods were used to predetermine sample sizes. We included all individuals with COVID-19 where sufficient material was available. No data were excluded from the analyses. Statistical details for each analysis are mentioned in each figure legend or in the respective part of the text. Single-nucleus RNA Sequencing (snRNA-seq), proteomics, and immunohistochemical stainings were performed in a blinded fashion. Evaluation of histological and immunohistochemical staining was performed separately by at least two independent (neuro-)pathologists. Histological staining, and immunohistochemistry were replicated at least once. The representative images shown were adjusted in brightness and contrast to different degrees (depending on the need resulting from the range of brightness and contrast of the raw images) in Adobe Photoshop or Adobe Illustrator, and for these cases raw image files are publicly available and deposited publicly at Zenodo (doi: 10.5281/zenodo.7524299). The experiments were not randomized.

## Data Availability

All data produced in the present study are available upon reasonable request to the authors.

## Acknowledgements

We gratefully thank F. Egelhofer, R. Koll, P. Matylewski, V. Wolf, S. Meier for excellent technical assistance and advice. The authors are most grateful to the individuals with COVD-19 and their relatives for consenting to autopsy and subsequent research, which were facilitated by the Biobank of the Department of Neuropathology, Charité–Universitätsmedizin Berlin. Cartoon images were partially created with Biorender.com. This work was supported by the Deutsche Forschungsgemeinschaft (DFG, German Research Foundation) under Germany’s Excellence Strategy EXC-2049-390688087, as well as SFB TRR 167 and HE 3130/6-1 to F.L.H., CRC130 P17 to H.R. and A.E.H., HA 5354/10-1 to A.E.H. and RA 2491/1-1 to H.R.; by the German Center for Neurodegenerative Diseases (DZNE) Berlin, by the European Union (PHAGO, 115976; Innovative Medicines Initiative-2; FP7-PEOPLE-2012-ITN: NeuroKine) and by the Ministry for Science and Education of Lower Saxony through the program ‘Niedersächsisches Vorab’ to J.F.

## Conflicts of interest

The authors declare no conflict of interests.

## Notes

### Competing Interest Statement

The authors have declared no competing interest.

### Funding Statement

This work was supported by the Deutsche Forschungsgemeinschaft (DFG, German Research Foundation) under Germanys Excellence Strategy EXC-2049-390688087, as well as SFB TRR 167 and HE 3130/6-1 to F.L.H., CRC130 P17 to H.R. and A.E.H., HA 5354/10-1 to A.E.H. and RA 2491/1-1 to H.R.; by the German Center for Neurodegenerative Diseases (DZNE) Berlin, by the European Union (PHAGO, 115976; Innovative Medicines Initiative-2; FP7-PEOPLE-2012-ITN: NeuroKine) and by the Ministry for Science and Education of Lower Saxony through the program Niedersachsisches Vorab to J.F.

### Author Declarations

Ethics commitee of Charite Universitymedicine Berlin Chariteplatz 1, 10117 Berlin (Germany) gave ethical approval for this study under the following registration numbers: EA1/144/13, EA2/066/20, EA1/075/19 and EA1/076/20.

