## Supplementary Figures for "The central nervous system’s proteogenomic and spatial imprint upon systemic viral infections with SARS-CoV-2"

#### Supplementary Figure 1

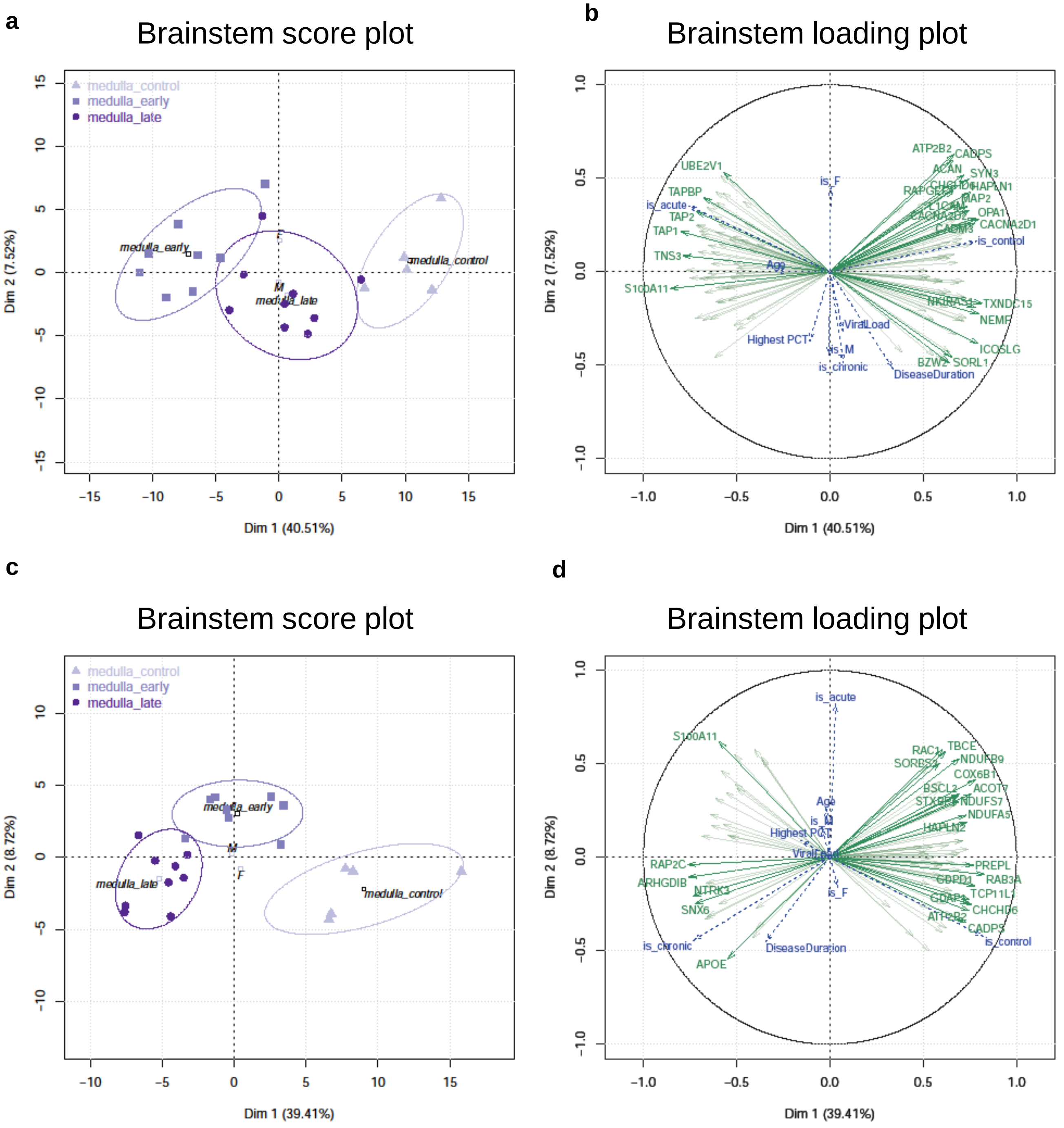

### Supplementary Figure 2

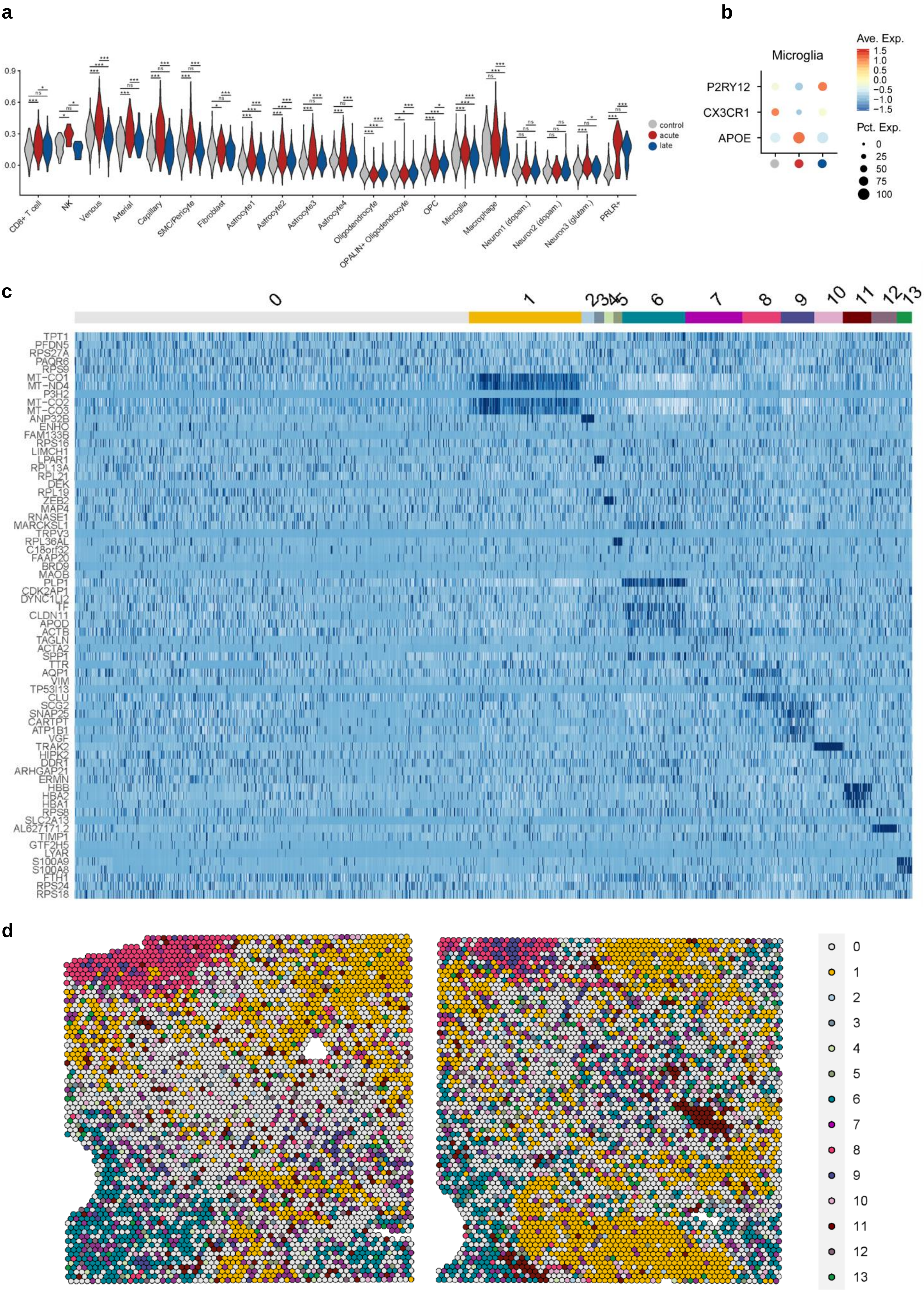

#### Supplementary Figure 3

**a**

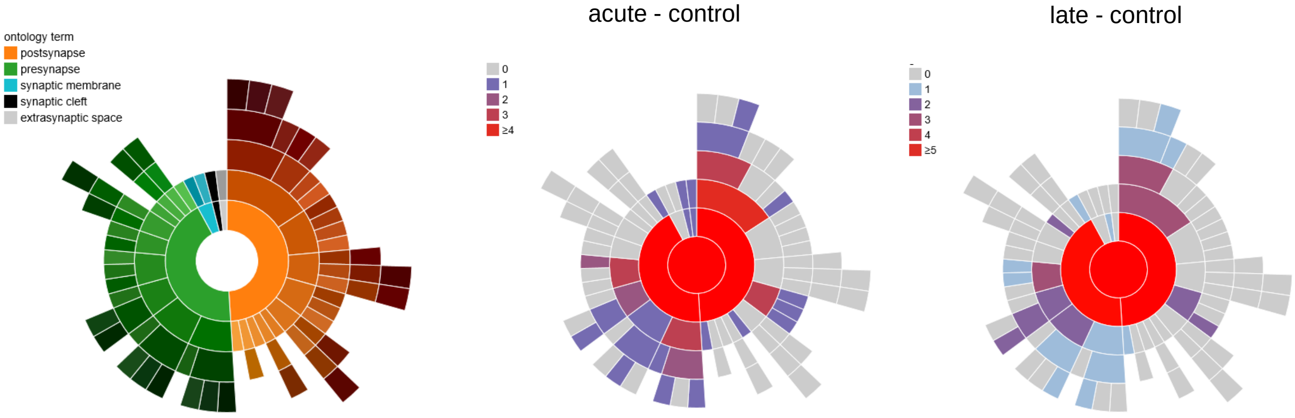


**b**

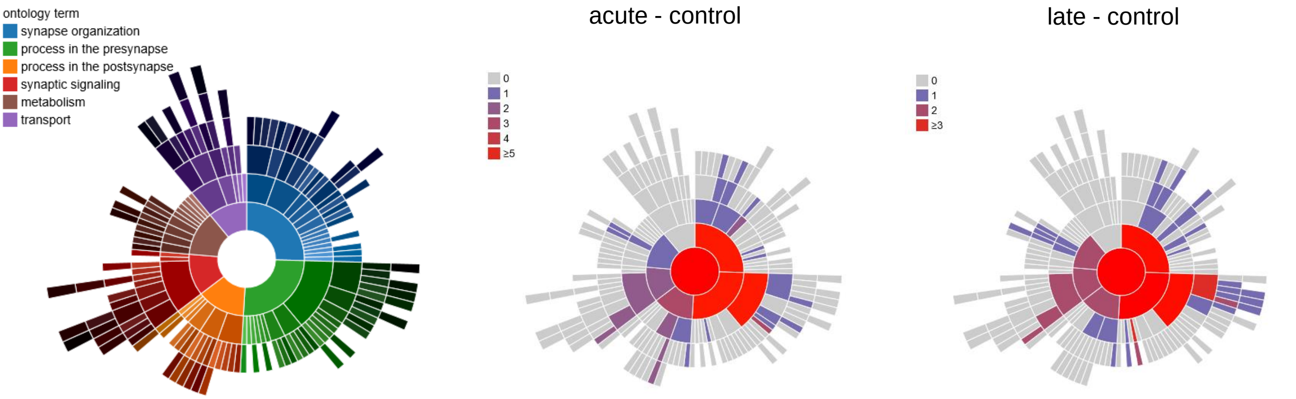


**c**

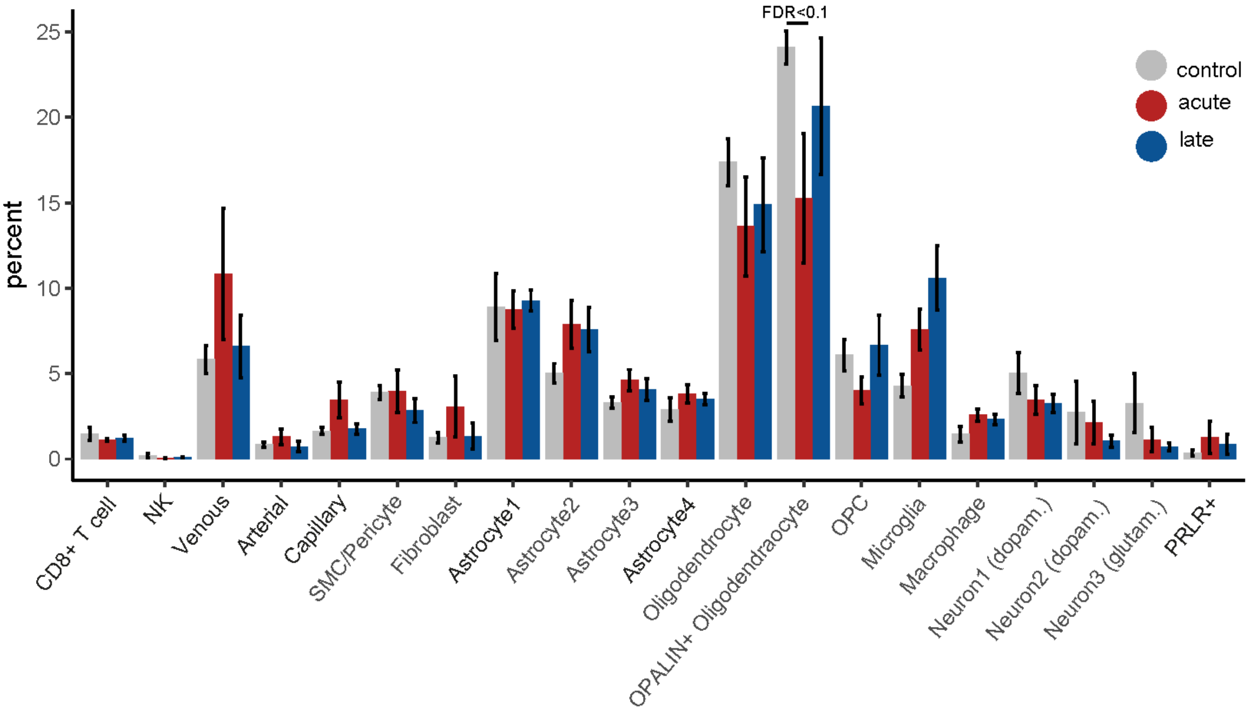

#### Supplementary Figure 4

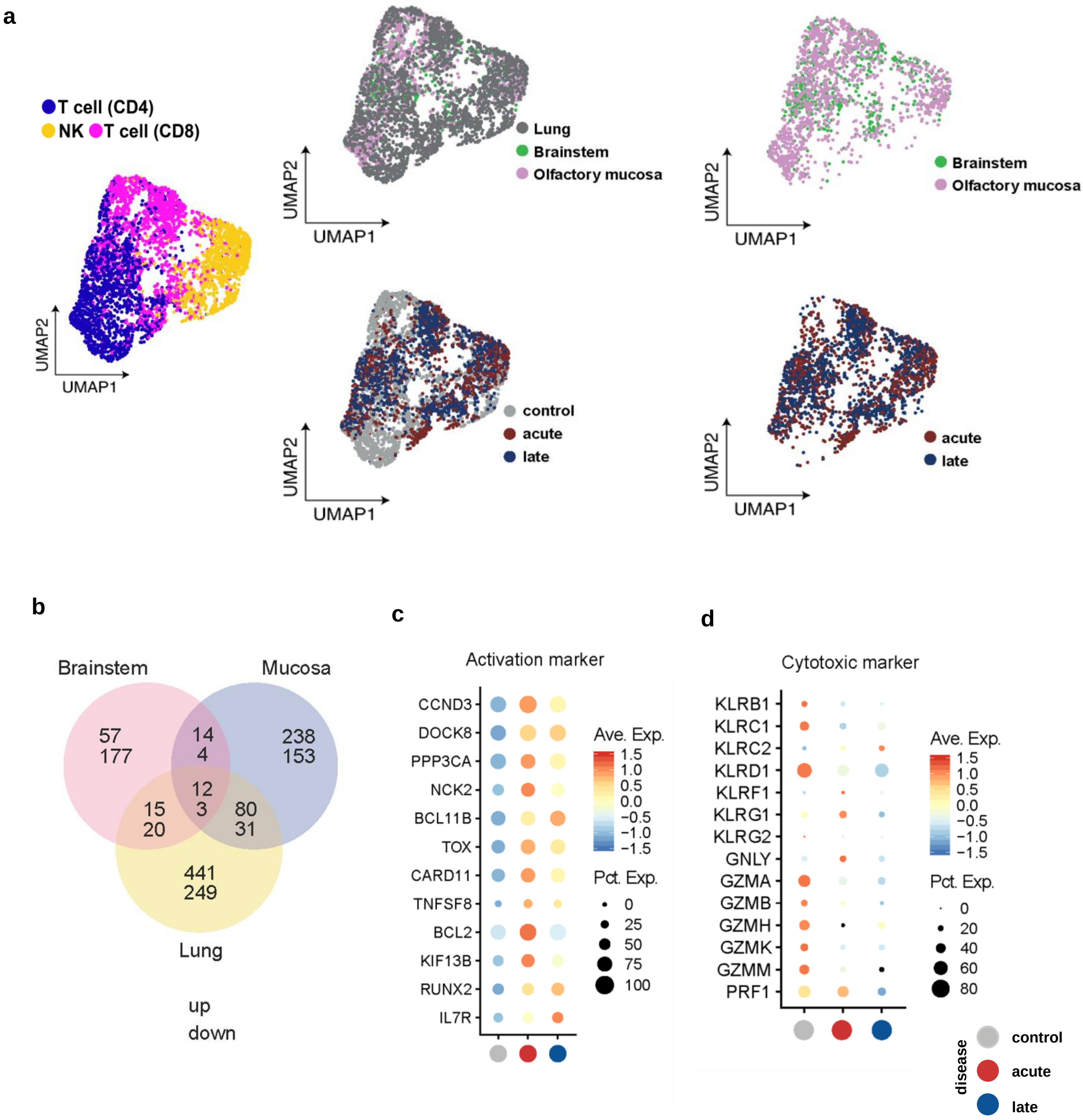

#### Supplementary Figure 5

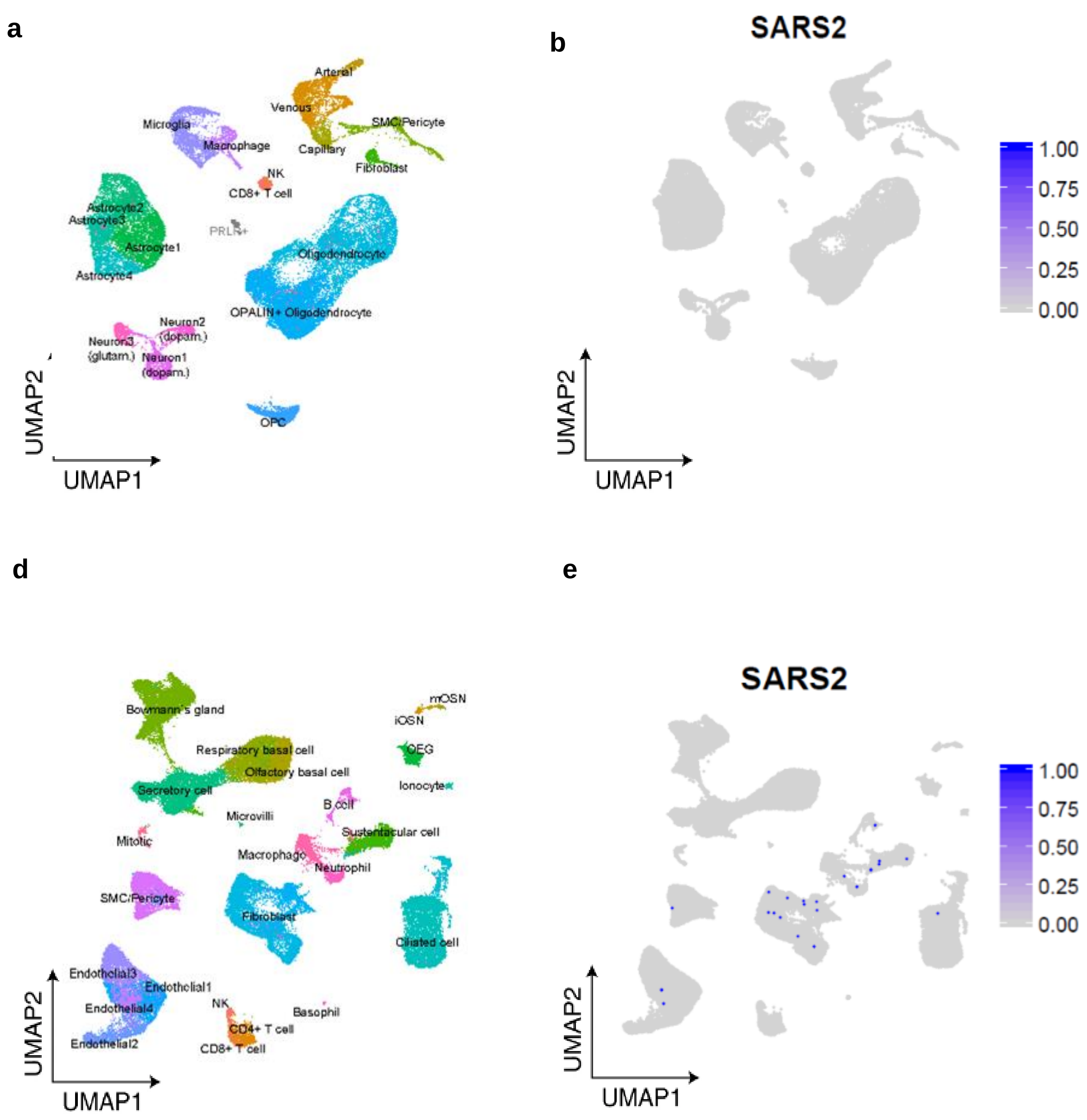

### Supplementary Figure 6

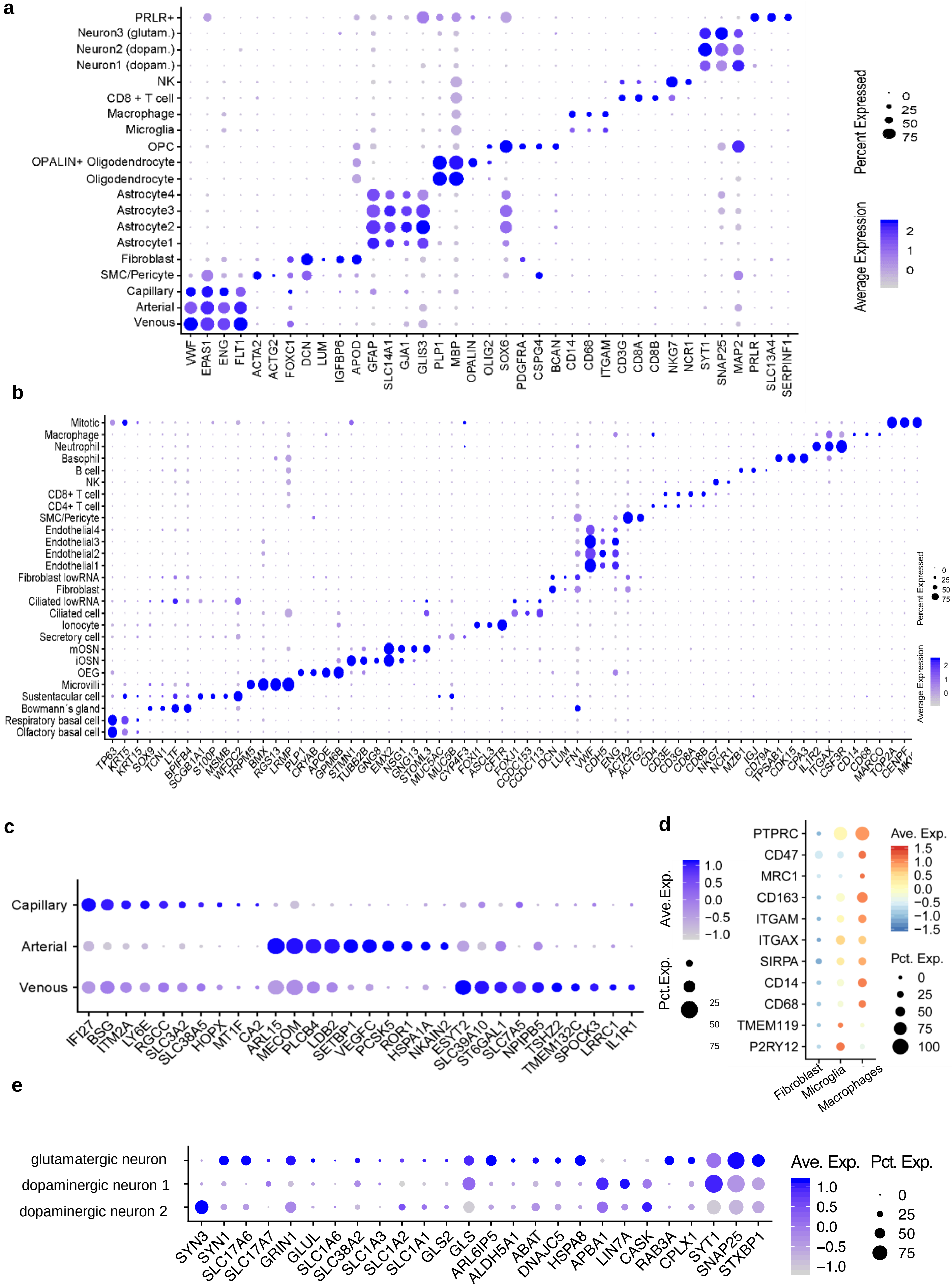
